# A novel framework for assessing causal effect of microbiome on health: long-term antibiotic usage as an instrument

**DOI:** 10.1101/2023.09.20.23295831

**Authors:** Nele Taba, Krista Fischer, Estonian Biobank research team, Elin Org, Oliver Aasmets

## Abstract

Assessing causality is undoubtedly one of the key questions in microbiome studies for the upcoming years. Since randomised trials in human subjects are often unethical or difficult to pursue, analytical methods to derive causal effects from observational data deserve attention. As simple covariate adjustment is not likely to account for all potential confounders, the idea of instrumental variable (IV) analysis is worth exploiting. Here we propose a novel framework of antibiotic instrumental variable regression (AB-IVR) for estimating the causal relationships between microbiome and various diseases. We rely on the recent studies showing that antibiotic treatment has a cumulative long-term effect on the microbiome, resulting in individuals with higher antibiotic usage to have a more perturbed microbiome. We apply the AB-IVR method on the Estonian Biobank data and show that the microbiome has a causal role in numerous diseases including migraine, depression and irritable bowel syndrome. We show with a plethora of sensitivity analyses that the identified causal effects are robust, and propose ways for further methodological developments.

## Introduction

Human microbiome studies have demonstrated that the gut microbiome can be affected by various exposures such as diet and medications^1,2^ and changes in the gut microbiome composition have been associated with the prevalence or susceptibility to complex diseases such as type 2 diabetes^3–5^, Crohn’s disease^6^ and different cancers^7,8^. However, with the primary interest in disease associations, determining the causal effects has remained challenging, as there are several potential confounders (dietary choices, other behavioural and environmental exposures) that may affect microbiome and their disease outcomes via different pathways. Experimental studies, such as utilising faecal microbiota transplantation, have proven a causal mechanistic link between treatment, microbiome and health^9^, but to date the methods for determining such causal associations noninvasively are limited^10^. An alternative would be to attempt causal effect estimation based on observational cohort data - using Instrumental Variable (IV) method to avoid biases due to unobserved confounding. One example of such an approach is Mendelian Randomization (MR), where a genetic variable acts as an instrument (affecting only the exposure variables, thus microbiota, but not directly the outcome or the confounders). Successful examples include studies that have shown the causal role of several taxa on cancers^11^, inflammatory bowel diseases^12^ and depression^13^. However, MR is limited by the lack of valid genetic instruments - this might remain an issue even with larger sample sizes, since microbiome is shown to be with low heritability^14,15^. To fill this void, we propose a new analytical approach to assess the causal role of microbiome on health: antibiotic instrumental variable regression (AB-IVR), using the long-term history of antibiotic usage (hAB) as an instrument.

Antibiotic (AB) usage is shown to affect microbiome in the long term in a cumulative manner, whereby larger number of antibiotics prescribed in the past results in a more perturbed microbiome composition^2,16^. This means that there are on average consistent differences in microbiome between the individuals with higher and lower long-term antibiotics usage. Importantly, this long-term usage effect is already evident in subjects who have taken only 3-5 courses of AB in the last 10 years before the sample collection. Such antibiotic consumption is highly common in the general population. This, and the fact that antibiotics are expected to affect health primarily via microbiome (directly eliminating the disease-causing bacteria, but also resulting in a general dysbiosis as a “collateral damage”) and not via any other pathways, made us ponder whether hAB - measured as number or antibiotics prescribed during 10 years prior to microbiome sampling - can serve as a natural experiment randomising individuals to have more or less perturbed microbiome. Thus, we propose that by using hAB as an instrument, we can compare the disease incidence between groups that have consistent long-term differences in microbiome, and by this assess the causal role of microbiome in disease. Of note, the primary focus of the AB-IVR method is on diseases, which itself are not treated with antibiotics. We consider the AB-IVR method as an intermediate option between the observational studies and randomised trials: we aim to gather more information compared to observational studies, but still do not see it as an equivalent alternative to the randomised trials.

Next sections introduce the AB-IVR methodology and its usage on the Estonian Biobank samples. Firstly, we introduce the method as a Two-Sample Two-Stage Least Squares (TSTSLS) procedure and the corresponding sample sets that we utilise for estimating the causal effects. Next, we introduce the concept of long-term antibiotics usage as an instrument. We discuss model assumptions and limitations that can bias the results such as the scope of the estimable effects, feedback loops arising from the bidirectional microbiome-disease crosstalk and effects of general health behaviour. Lastly, we assess the causal role of microbiome in several common diseases, test the validity of the findings through a series of sensitivity analyses and give insights for future developments. The AB-IVR method showed that microbiome has a causal effect for several common diseases such as irritable bowel syndrome, migraine and depression, and we believe that it opens up new possibilities for causal discovery in the human microbiome field.

## Results

### Study overview and data selection

For inferring the causal effect of microbiome on disease we utilised the method of Two-Sample Two-Stage Least Squares (TSTSLS), which is a special case of instrumental variable regression. In our case, the history of antibiotics usage (hAB) serves as the instrumental variable, hence the term antibiotic instrumental variable regression, AB-IVR (**Fig. 1A**). The general idea of the method is that when AB usage cannot realistically have any direct effect on the disease of interest, the correlation of AB usage with the disease incidence can only arise via mediation by microbiome. Thus the total effect of hAB on the disease outcome can be decomposed into the effect of hAB on the microbiome and the causal effect of the microbiome on the outcome. As the total effect and the effect of hAB on the microbiome are estimable, also the causal effect of the microbiome becomes identifiable.

**Figure 1.**
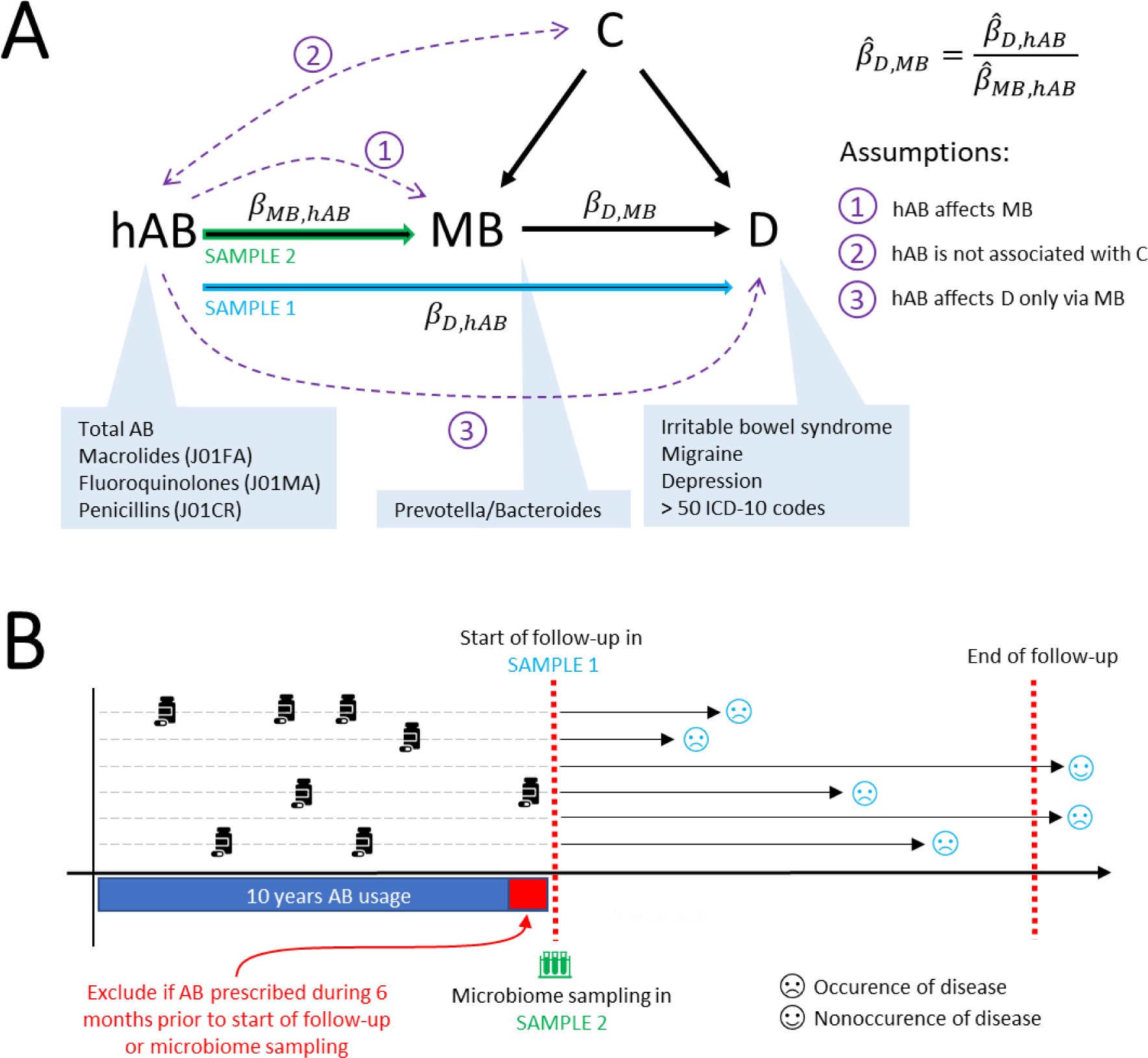
Graphical description of the study design. Upper panel **(A)** illustrates the instrumental variable regression and corresponding assumptions schematically in the context of our study. hAB - history of antibiotic usage; MB - microbiome; D - disease; C - confounder. Lower panel **(B)** illustrates the samples and data used. Sample 1 refers to the sample where disease follow-up data is recorded, EstBB in our case. Sample 2 refers to the sample where the microbiome is assessed, EstMB in our case. We recorded the AB usage as the number of AB prescribed (total and AB subgroups) during the 10-year period preceding the start of follow up in Sample 1 and microbiome sampling in Sample 2, whereas in both samples the individuals receiving AB during the last 6 months of the aforementioned period are excluded. Sample 1 was followed up for incident outcomes of 56 common diseases from 01.01.2015 until 31.12.2022, whereafter observations are right censored irrespective of the future outcomes. In Sample 2 the microbiome was assessed on an arbitrary moment between November 2017 to July 2020. The causal effect of MB on disease (β_*D,MB*_) in the two-sample setting is estimated as the ratio of the effect of AB-usage on disease (β_*D,hAB*_) in Sample1 (EstBB) and the effect of AB-usage on MB (β_*MB,hAB*_) in Sample2 (EstMB).

In our study, the TSTSLS methodology relies on two independent datasets which are used for 1) estimating the effect of hAB on disease (β_**D,hAB**_) and 2) estimating the effect of hAB on the microbiome (β_**MB,hAB**_). The two effects are combined to evaluate the causal effect of microbiome on disease (β_**D,MB**_)(**Fig. 1A**). For estimating the effect of hAB on disease, we leveraged the electronic health records data available for the Estonian Biobank (EstBB) participants (N > 210 000)^17^. As prevalent diseases can have an effect on the microbiome and potentially create a feedback loop, which biases the results, we analysed the effect of previous antibiotics usage on the incident diseases instead (See Methods). To estimate the effect of hAB on the microbiome, we used Estonian Microbiome (EstMB, a subcohort of EstBB) cohort (N > 2500) samples with shotgun metagenomics data available^2^.

### History of antibiotics usage as a valid instrument

Estimating the causal effects by instrumental variable regression analysis relies on several assumptions. Firstly, the hAB must be associated with the gut microbiome (**Fig. 1A**). We and others have previously shown that hAB is associated with the microbiome composition additively, meaning that higher hAB is associated with larger changes in the microbiome composition^2,16^. Here, we focused our analysis on the *Prevotella*/*Bacteroides* ratio (P/B ratio) that represents a summarised state of microbiome by reflecting the enterotype composition^18^. We confirm the additive effect of total hAB on P/B ratio and show that it also holds for different classes of antibiotics, namely macrolides (ATC code J01FA), penicillins (J01CR) and fluoroquinolones (J01MA), which we will use for the sensitivity analysis (**Fig. 2A**). It must be highlighted that the different antibiotics classes are largely uncorrelated in their usage and their long-term effects on the microbiome composition differ (**Fig. 2B**, **Fig. 2C**).

**Figure 2.**
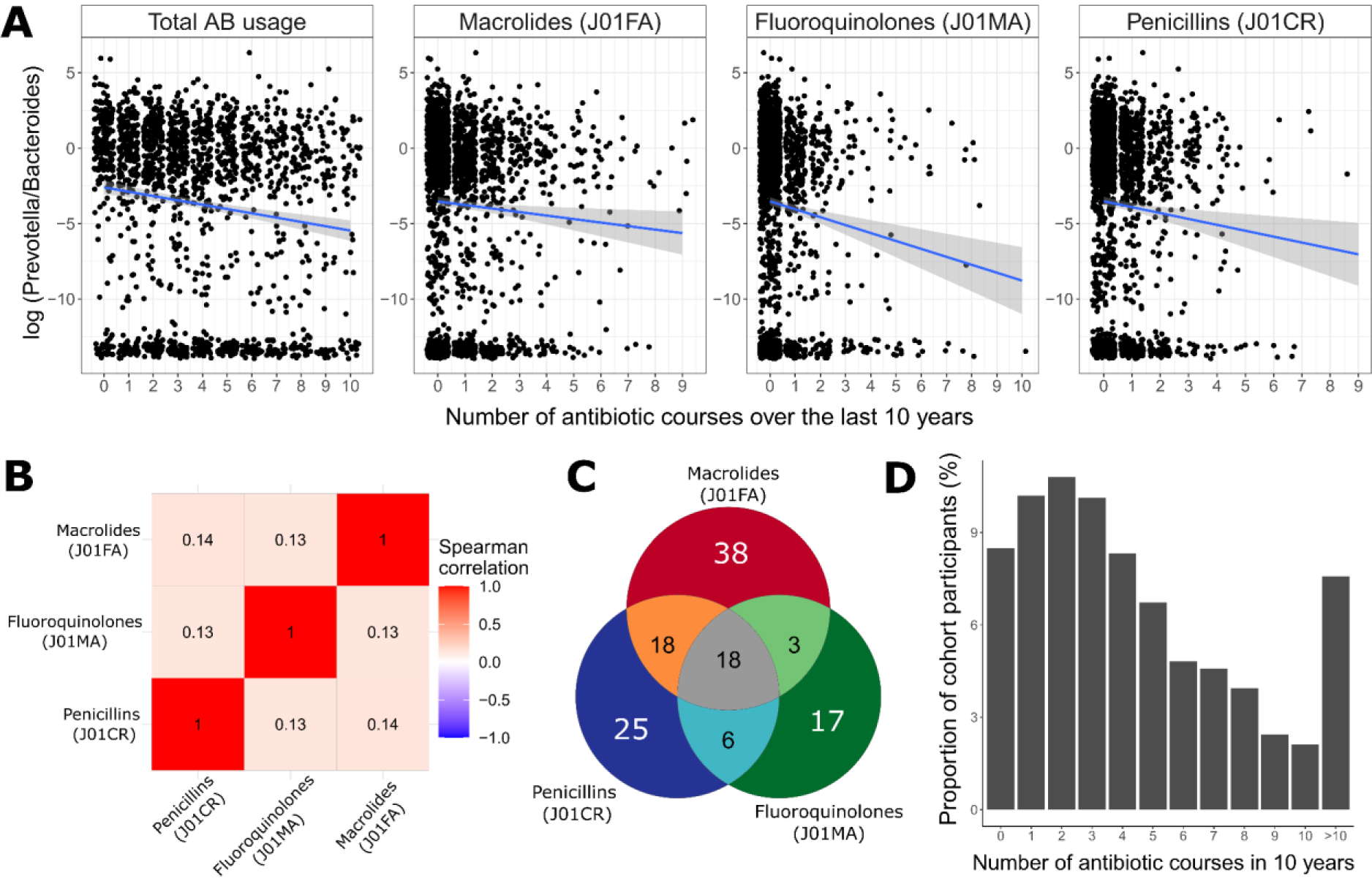
Antibiotic usage in the EstMB cohort. Panel **A** shows the association between the number of different antibiotics used during the last 10 years before the sample collection and *Prevotella-Bacteroides* ratio. The total AB usage and usage of all AB subclasses was strongly associated with *Prevotella/Bacteroides* ratio (combined AB usage p=2.04e-8, macrolides p=0.0192, fluoroquinolones p=3.03e-5, and penicillins p=0.0026). Panel **B** shows the Spearman correlation between the number of different antibiotics classes used during the last 10 years before the sample collection. Panel **C** shows the unique and shared hits of the univariate analyses associating the antibiotics usage history with the abundance of microbial species. Panel **D** shows the proportion of cohort participants by the number of antibiotics used during the past 10 years.

Secondly, the usage of antibiotics and the disease cannot have any common cause. This assumption can be easily violated when prevalent disease cases are analysed. Namely, the disease could for example weaken the immune system, which would in turn result in higher antibiotics consumption, often referred to as a “feedback loop”. We address this issue by analysing only incident disease cases that are observed during the follow-up period (thus after the period where AB usage was recorded). As a sensitivity analysis we additionally analyse disease cases that have been diagnosed more than 5 years after the sample collection, to eliminate the possibility of prevalent diseases that have not yet been diagnosed, but are already present and affecting the microbiome composition. Violation of the second assumption can also occur when subjects of poorer health are studied, since these individuals are more likely to take a larger number of antibiotics and develop further comorbidities. For example, elderly individuals are likely to have weaker immunity and therefore take a larger number of antibiotics, but at the same time they are at higher risk of various comorbidities. Thus an overall frailty level could act as a confounder. To address this concern, we focus our analysis only on younger people (ages 23-50), who have taken up to 5 courses of antibiotics during 10 years prior to microbiome sampling or start of follow-up (See Methods, **Fig. 1**). The lower limit for age refers to the minimum age in the EstMB cohort. The chosen threshold for the number of antibiotics courses is highly common in the population, thus it is meant to represent the healthier part of the population (**Fig. 2D**), allowing us to exclude individuals with chronic and extreme AB usage. As sensitivity analyses, we study older people, extending the age range to 89 (refers to the maximum age in EstMB cohort), and usage of antibiotics up to 10 courses. Thirdly, antibiotics cannot have a direct effect on the disease that is not mediated by the microbiome composition. Although some authors have indicated that antibiotics can have such microbiome-independent effects, the evidence is currently weak and we consider that most of the effect is based on altering the microbiome composition^19^.

### Causal role of the microbiome on diverse disease groups confirmed with the novel AB-IVR framework

We applied the AB-IVR methodology to assess whether the microbiome has a causal role for the development of 56 conditions across diverse disease groups. We focused primarily on common chronic diseases and cancers where antibiotics are not used as a treatment and which had at least 50 incident cases. Due to the broad selection of diseases, we did not apply any disease specific inclusion and exclusion criteria, but future studies should take the disease specificity into account. Antibiotics have a complex effect on MB via altering the abundances of several members of the composition simultaneously. Therefore, when the analysis is based on general hAB, various taxa are altered simultaneously and thus it is not possible to imply which specific species or genera are the culprit in disease formation. This is a limitation of our methodology and remains a challenge to be solved by future improvements. Therefore, instead of using the data of specific species or genera to characterise the changes in the microbiome, we analysed the effect of *Prevotella-Bacteroides* (P/B) ratio, which reflects the general state of the microbiome.

In total, the microbiome was identified to have a causal effect on 23 diseases (FDR <= 0.05) (**Fig. 3A**, **Supplementary Fig. 1, Supplementary Table 1**). These diseases include cardiometabolic diseases like cardiac arrhythmias, intestinal diseases like irritable bowel syndrome and gastro-esophageal reflux disease, skin diseases like atopic dermatitis and acne, and several mental disorders like anxiety disorders and depression. The causal estimates are largest for lactose intolerance, migraine and irritable bowel syndrome with the dominance of *Prevotella* defending against the disease progression. Notably, we did not identify the causal effect of the microbiome on any of the cancers studied, which is likely due to excluding older individuals from the analysis.

**Figure 3.**
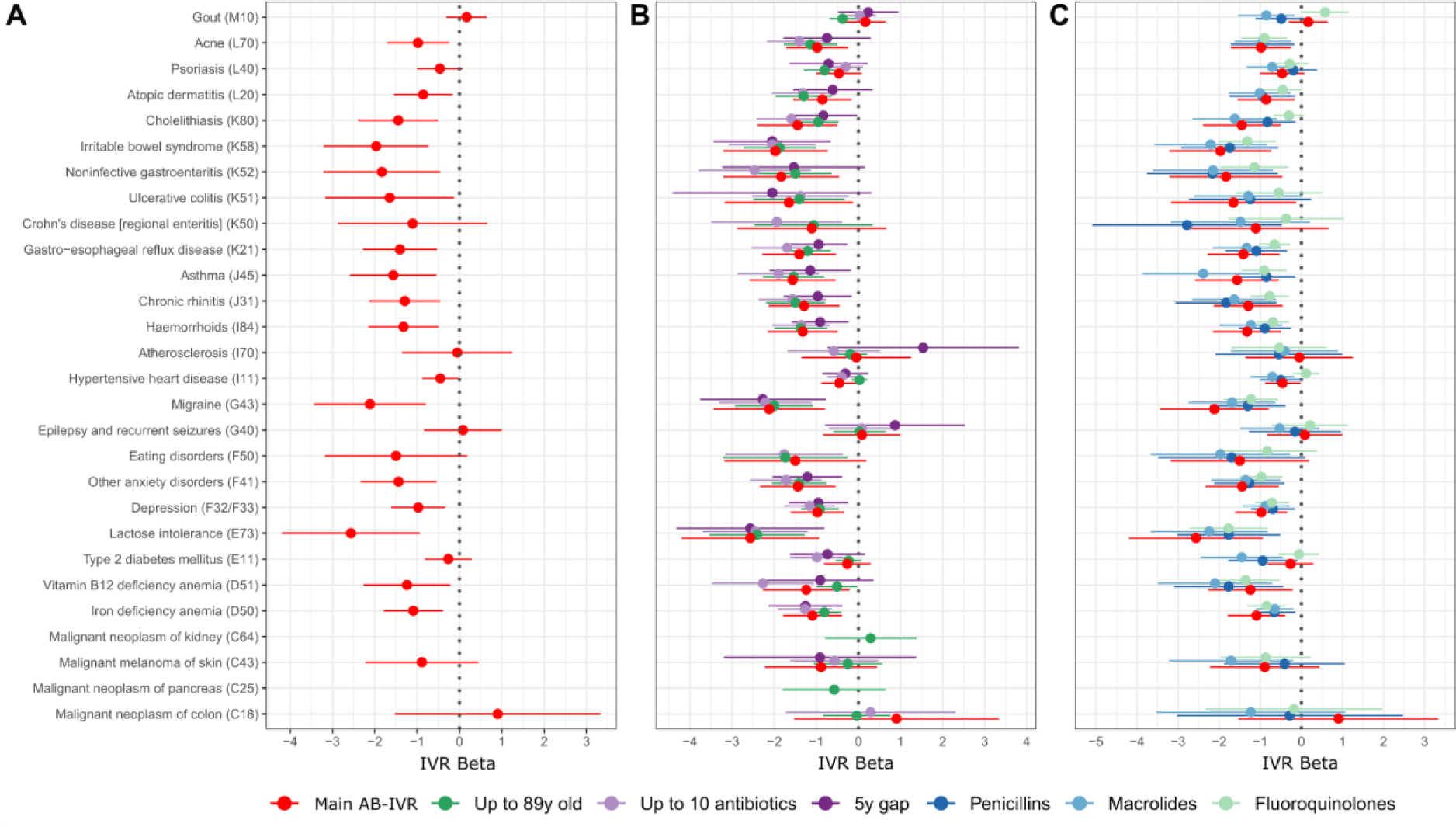
Results of the main AB-IVR analysis (A) and sensitivity analyses. **(B,C).** On panels **A**,**B**, and **C** the causal effect estimate of *Prevotella*/*Bacteroides* ratio on a selection of diseases is presented (full results comprising all diseases analysed can be viewed in **Supplementary Tables 1, 3-5, and 6-8**). In panel **A** the age is filtered as 23-50, maximum number of AB prescribed is five, and minimum number of cases per disease is 50. Panel **B** represents the sensitivity analyses where sample formation varies, whereas the main analysis is depicted for comparison in red. Green shows the effect estimates, when age is filtered as 23-89; light-purple shows the effect estimates, when the maximum number of AB prescribed is 10; dark-purple describes the scenario where the first five years of incidence after the start of follow-up is considered as prevalent disease. Panel **C** represents the sensitivity analyses where information regarding subclasses of AB were used instead of the total amount of AB prescribed, whereas other settings were identical to the main analysis. Red corresponds to the main analysis (same as **A**), dark blue corresponds to the class of penicillins (J01CR), light-blue corresponds to the class of macrolides (J01FA) and light-green corresponds to the class of fluoroquinolones (J01MA).

### Sensitivity analysis and assessment of different antibiotics as instruments

We carried out several sensitivity analyses to assess the validity of the model estimates. Firstly, we simulated random binary variables with different event probabilities to see how our approach behaves in a scenario where no effect is expected. Indeed, the method did not identify any effects and manages to control the false discovery rate perfectly (**Supplementary Table 2**). Next, we focused on different scenarios which would test the model behaviour when the study population includes subjects who are more likely to have health complications, thus testing the second model assumption. For that we considered subjects up to 89 years old (refers to the maximum age in EstMB cohort), subjects that had been prescribed antibiotics up to 10 times during 10 years prior to microbiome sample collection or start of follow-up, and considering a 5 year “gap” in defining the incident cases after the start of follow-up (**Fig. 3B, Supplementary Fig. 2, Supplementary Tables 3, 4, 5**). The reasoning behind the chosen sensitivity analyses is discussed in detail in the Methods. We observed no major differences in the estimated causal effects for any of the tested scenarios. Most notably, the causal estimates for diseases that showed strongest effects in the primary analysis remained largely untouched by the scenarios tested. This includes diseases like migraine, irritable bowel syndrome, chronic rhinitis and depression. Lastly, we carried out sensitivity analysis to test the performance of different antibiotics subclasses as instruments (**Fig. 3C, Supplementary Tables 6, 7, 8**). Again, the results remained similar and the previously highlighted causal estimates remained largely unchanged and independent of the chosen instrument. Since different antibiotics classes are largely uncorrelated in their usage and their long-term effects on the microbiome composition differs (**Fig. 2B**, **Fig. 2C**), the differences between the effect estimates can originate from the varying targets of antibiotic subclasses and can refer to divergent patterns in influenced subcommunities. This might be the case for gout where using macrolides and fluoroquinolones as instruments results in causal effect estimates with opposite directions, but with the total antibiotics usage as an instrument, no causal effect is identified (**Supplementary Fig. 3**). However, the overall similarity of the estimates for most of the diseases confirms the underlying causal effect of the microbiome as a whole. Lastly, since the logarithm of the P/B ratio is with a bimodal distribution, we additionally performed an analysis where the P/B ratio was inverse normal transformed. However, we did not observe any notable discrepancies compared to the main analysis (**Supplementary Fig. 1, Supplementary Table 9**).

## Discussion

We introduce and demonstrate an analytical approach that expands the causal inference toolbox for microbiome studies by using long-term antibiotics usage as an instrument in an instrumental variable regression setting. We show that the history of antibiotics usage can be a valid instrument and that the microbiome has a causal role for several diseases, such as migraine, irritable bowel syndrome, depression and many others.

Establishing the causal role of microbiome in complex diseases can involve different levels of evidence from the identification of disease associations and faecal microbiota transplantations (FMT) to understanding the molecular mechanisms that produce the phenotype^20^. While the established knowledge about the mechanisms of microbial strains or microbial metabolites remains scarce, computational approaches and FMT studies have been used to show the causal role of microbiome in several conditions^20^. In line with our findings, Mendelian randomization studies have identified the causal role of microbial taxa for migraine^21^, irritable bowel syndrome^22^ and depression^13,23^. Nevertheless, MR studies have indicated the causal role of microbiome in type 2 diabetes^23^, psoriasis^24^, inflammatory bowel diseases^12^ and heart failure^25^, which was inconsistent with our findings. However, these inconsistencies can be expected as our approach focuses on the general effects of the microbiome composition, which is more in line with how the FMT studies can inform causality. Then again, FMT studies are coupled with challenges such as screening and processing the FMT material and selection of the donor and the mode of delivery^26^. Additionally, the gut microbiome and lifestyle of the recipient can significantly alter the success of the FMT^26^. Therefore, although FMT has been shown to transmit or alleviate hypertension^27^, IBD^28^ and insulin sensitivity^29,30^, the results can be conflicting^28,31^. Thus, we believe that in the quest for establishing the causal effects, our approach can provide an additional layer of evidence that combines features from the MR and FMT approaches.

Antibiotics are meant to kill or inhibit the growth of bacteria and are commonly directly or indirectly effective against several members of the microbial community^32^. Therefore, using long term usage of antibiotics as an instrument allows us to estimate the causal effect of the disturbed part of the community. This brings along two limitations for the proposed methodology: we cannot identify the effects of single members of the community; and we cannot rule out the potential causal effects of the members that are unaffected by the antibiotics used. Thus, the microbiome can still be causal for the development of diseases for which our proposed methodology did not show a causal effect. Tackling these limitations opens up several directions for future development. Since different antibiotic subgroups are intended to have an effect on different bacteria, one can use or design instruments that are specific to certain subcommunities or taxa to estimate the causal effects of interest. Also, several instruments such as long-term usage of different antibiotic subclasses and other drugs with persistent effects on microbiome can be combined and used simultaneously, which in turn opens the opportunity to utilise various sensitivity analyses that are developed for 2-sample Mendelian Randomization framework. For example, consumption of numerous host-targeted drugs are known to affect the microbiome^16,33^ and antidepressants have also been shown to have long term effects^34^, but as microbiome-related research is currently rapidly evolving, many more suitable instruments, not limited to medications, are likely emerging in the near future.

Here, we showed in principle how long-term effects of antibiotics can be used to identify the causal role of the microbiome in a large set of common diseases. However, certain limitations must be kept in mind while interpreting the results for a specific outcome. We did not focus separately on any single disease, thus future studies should consider disease-specific inclusion-exclusion criteria when designing the study. An important consideration concerns the role of general health behaviour. It is possible that the overall awareness and interest in one’s health promotes the usage of antibiotics, which also indirectly changes the disease risks. Variables such as gender and education might entail general health behaviour and consequently simultaneously affect AB-usage and health-outcomes (eg. individuals with higher education might seek more help from health-care professionals (thus have different AB-consumption habits), have better eating habits, smoke less, be more physically active etc). Therefore we advise any future studies using AB-IVR framework to account for such possible confounders. Lastly, the amount of antibiotic consumption is highly variable in different populations and antibiotic consumption in Estonia is among the lowest in Europe^35^. Thus, the results need to be validated in different populations to account for the differences in overall burden of antibiotics usage.

We demonstrated a novel AB-IVR approach by assessing the effect of *Prevotella/Bacteroides* ratio on a large set of diseases. The results confirmed the utility of the introduced method and we believe it has large potential to become a new widely used framework that allows to assess analytically the causal effect of microbiome on health. Further research with disease-specific inclusion-exclusion criteria is warranted for drawing conclusions about specific health-effects and several exciting future development options arised.

## Methods

### Sample description

The proposed method combines the information from two different samples, whereas the association between the instrument and the exposure is analysed in one sample and the association between the instrument and the outcome in another sample (**Fig. 1**). In our case the association between antibiotics usage and microbiome is assessed in the Estonian Microbiome Cohort^2^ and association between antibiotics usage and diseases in the Estonian Biobank data^17^.

The Estonian Biobank (EstBB) is a volunteer-based population cohort initiated in 1999 that currently includes over 210,000 genotyped adults (≥ 18 years old) across Estonia. Estonian Microbiome Cohort (EstMB) was initiated in 2017 when more than 2500 EstBB participants, who joined the EstBB at least 10 years before, provided stool, oral, and blood samples. The EstMB cohort, microbiome sample collection, stool bacterial DNA extraction and shotgun metagenomic sequencing are described in detail in Aasmets & Krigul *et al.*^2^. For the current project, taxonomic profiling of the gut microbiome was carried out using *MetaPhlAn3 tool*^36^. The *Prevotella-Bacteroides* ratio, which we considered as the primary indicator of the microbiome inter-individual variability in our analysis, was calculated on the *genus* level after imputing zeros with a pseudocount equal to half of the minimal non-zero relative abundance observed in the data. For both, EstMB and EstBB one of the major advantages is the possibility to use electronic health records (EHR) data and follow the participants’ health both retrospectively and prospectively. Using EHR, we obtained the data for the history of antibiotics usage and the disease incidence. The EstMB participants were removed from the EstBB sample to ensure that the two samples are independent.

### Start of follow-up

An important aspect in the following data-processing is the moment when MB is collected in Sample 2 (EstMB) and the follow-up starts in Sample 1 (EstBB) (**Fig. 1**). Of note, since the proposed methodology uses two independent samples, the MB collection and start of follow-up do not need to be at the same moment of time. In the EstMB cohort the time of MB sample collection varies between individuals, whereas in the EstBB cohort the start of follow-up is set to 1st January 2015 for all individuals. The reason for the latter is that the history of antibiotics usage is not properly recorded before 2005 in the electronic health records and the chosen cut-off date allows us to analyse the antibiotics consumption during a 10-year period prior to follow-up in the EstBB cohort. As we were interested in the long-term effects rather than the extreme alterations that occur due to recent antibiotics usage, we excluded the individuals who had been prescribed AB during the six months preceding either the start of MB sample collection in EstMB or start of follow-up in EstBB.

### Defining antibiotics usage and incident diseases

To characterise the long-term antibiotics usage, we quantified the number of antibiotics treatments prescribed in the period between the last 6 months and last 10 years before the microbiome sample collection or start of follow-up. All drugs with the Anatomical Therapeutic Chemical (ATC) classification code J01* were considered as antibiotics. We chose the 10-year period similar to Aasmets & Krigul *et al.*^2^, where we showed a cumulative long-term effect of antibiotics usage. Additionally, we considered separately the history of usage of penicillins (ATC code J01CR), macrolides (J01FA) and fluoroquinolones (J01MA) since these classes have the strongest long-term effects on gut microbiome (Aasmets *et al., unpublished data*). We restricted the analysis to incident diseases, which we defined as the first occurrence of an ICD-10 code of interest in the EHR after the start of follow-up (1st January 2015) until the end of follow-up (31st December 2022). We focused primarily on the chronic common diseases and cancers with at least 50 incident cases in the EstBB cohort. Summary of the selected diseases is shown in **Supplementary Table 1**. Furthermore, we excluded from any analysis of an incident disease the prevalent cases for the corresponding disease. We did not use any other disease-specific exclusion criteria for the method demonstration. However, we do encourage to utilise disease-specific exclusion criteria when a more thorough analysis of a specific disease is of interest.

### Main method

For inferring the causal effect of MB on disease we utilised the method of Two-Sample Two-Stage Least Squares (TSTSLS), which is a special case of instrumental variable regression for the two-stage estimation in a two-sample setting. A detailed description of TSTSLS can be found elsewhere^37^. Rationale for using the two-sample setting is that the microbiome is often measured in smaller samples compared to incident disease outcomes available in the large biobank samples. Using TSTSLS allows us to leverage the information from such large samples. We denote in the following formulas the estimated effect of instrument (hAB) on exposure (MB) as β_*MB,hAB*_, instrument on outcome (D) as β_*D,hAB*_, and exposure on outcome, i.e. the causal relationship of interest, as β_*D,MB*_. We estimate β_*D,hAB*_ in the EstBB cohort (Sample1) using the logistic regression model and β_*MB,hAB*_ in the EstMB cohort (Sample2) using the linear regression model. Thereafter we calculate the effect-estimate for the causal relationship as a ratio of coefficients:

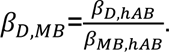

Further, we calculate the standard deviation for the estimated β_*D,MB*_ based on Pacini & Windmeijer^37^ as:

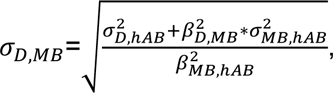

where 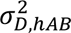 is the variance of 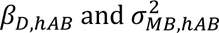 is the variance of β_*MB,hAB*_. We obtained the p-values based on z-scores, calculated as β_*D,MB*_/σ_*D,MB*_, and standard normal distribution. Further, we calculated the confidence intervals for the estimates as β_*D,MB*_ ± 1.96 ∗ σ_*D,MB*_.

To account for multiple testing, we used the Benjamini-Hochberg procedure to control for the false discovery rate (FDR). We set the significance threshold for FDR-adjusted p-value to 0.05.

### Model assumptions

The obtained β_*D,MB*_ is a valid causal estimate if the following assumptions are met^38^:

● AB usage history has an effect on MB;
● AB usage history is not associated with any of the confounders of the microbiome-disease association;
● There is no direct effect of AB usage history on incident disease outcomes outside of the pathway via microbiome.

Possible violations of these assumptions are described in the Results section.

### Sensitivity analyses

In the main analysis, we set the maximum number of antibiotics prescribed during the period of interest to 5, minimum age to 23 and maximum age to 50. The lower limit for age refers to the minimum age in the EstMB cohort. Subjects with impaired health might be more prone to comorbidities and higher consumption of antibiotics. Thus, the aim of the initial restriction of the age range and number of antibiotics used is to focus on the healthier part of the population to address potential confounding. However, we pursued several sensitivity analyses to assess the validity and robustness of the method:

1. We assessed the performance of the method in the setting, where we expect there to be no association. For that we created pseudo-variables indicating disease presence/absence for each individual in the EstBB sample by randomly sampling from binomial distribution with predefined probability. In total we created 39 pseudo-variables with disease-probabilities ranging from 2.5% to 97.5% (**Supplementary Table 2)**. The purpose of the analysis was to check whether the proportion of significant associations between randomly generated disease-variables and P/B ratio remains below the nominal alpha-level of 5%.
2. To account for the feedback-mechanism of the disease whereby the disease might actually already be prevalent for a period of time prior to diagnosis and could thus already have an effect on the MB, we pursued a sensitivity analysis where we considered the first 5 years of incidence after the start of follow-up as prevalent and therefore excluded such individuals from the analysis **(Supplementary Table 5)**. With such sensitivity analysis we aim to eliminate bias originating from the feedback mechanism of a disease that was already developing and affecting MB prior to the diagnosis;
3. We assessed the relationships between MB and diseases while using specific AB subgroups as instruments instead of total AB-usage. More specifically, we defined three additional variables as instruments based on the usage of AB with following ATC-codes: J01FA (macrolides) **(Supplementary Table 6)**, J01MA (fluoroquinolones) **(Supplementary Table 7)**, and J01CR (penicillins) **(Supplementary Table 8)**. These antibiotics classes were chosen since they have the strongest long-term effects on gut microbiome (Aasmets *et al., unpublished data*). All three variables were uncorrelated with each other (all Spearman correlations in EstMB and EstBB between the three AB-subgroups below 0.2 (**Fig. 2B**)), thus representing independent instruments. We assumed to see in general similar patterns of associations between MB and diseases, since a strong causal effect should not depend on the instrument used, provided that the instrument is valid. For this sub-analysis we excluded all the individuals who were prescribed any AB regardless of the ATC-code during 6 months prior to MB sample collection or start of follow-up. These analyses were carried out similarly to the main analysis with the maximum number of AB equal to 5 and age between 23 and 50.
4. Since the logarithm of the *Prevotella-Bacteroides* (P/B) ratio has a bimodal distribution, we additionally performed an analysis where we applied inverse normal transformation on the P/B ratio prior to analysis using the *RNOmni* R package^39^. **(Supplementary Table 9)**

## Data availability

The metagenomic data generated in this study have been deposited in the European Genome-Phenome Archive database (https://www.ebi.ac.uk/ega/) under accession code EGAS00001008448. The phenotype data contain sensitive information from healthcare registers and they are available under restricted access through the Estonian biobank upon submission of a research plan and signing a data transfer agreement. All data access to the Estonian Biobank must follow the informed consent regulations of the Estonian Committee on Bioethics and Human Research, which are clearly described in the Data Access section at https://genomics.ut.ee/en/content/estonian-biobank. A preliminary request for raw metagenome and phenotype data must first be submitted via the email address

All participants included in the EstBB cohort provided informed consent for the data and samples to be used for scientific purposes. All participants have joined the Estonian Biobank on a voluntary basis and have signed a broad consent form, which allows to receive participant’s personal and health data from national registries and databases. Rights of gene donors are regulated by Human Genes Research Act (HGRA) § 9 – Voluntary nature of gene donation (https://www.riigiteataja.ee/en/eli/ee/531102013003/consolide/ current). This study was approved by the Research Ethics Committee of the University of Tartu (approval No. 266/T10) and by the Estonian Committee on Bioethics and Human Research (Estonian Ministry of Social Affairs; approval No. 1.1-12/17 and 1.1-12/624).

## Acknowledgements

This study was funded by the European Union through the European Regional Development Fund Project No. 2014-2020.4.01.15-0012 GENTRANSMED. Data analysis was carried out in part in the High-Performance Computing Center of University of Tartu.

The authors would like to thank Mari-Liis Tammeorg, Kreete Lüll, Marili Palover, Anu Reigo, Neeme Tõnisson, Liis Leitsalu, Triinu Temberg and Esta Pintsaar for participating in the sample collection process of the Estonian Microbiome cohort. We thank Steven Smit, Rita Kreevan and Martin Tootsi for the DNA extraction process. We thank Reidar Andreson for bioinformatic support. We also thank all the EstBB study participants.

## Author Contributions

O.A., E.O. and K.F. conceptualised and supervised the study. O.A and N.T. designed the study and performed the data analysis. N.T. and O.A. interpreted the data and prepared the figures and wrote the manuscript. The EstBB Research Team provided the phenotype data. All authors read and approved the final paper.

## Supplementary figures

**Supplementary figure 1.**
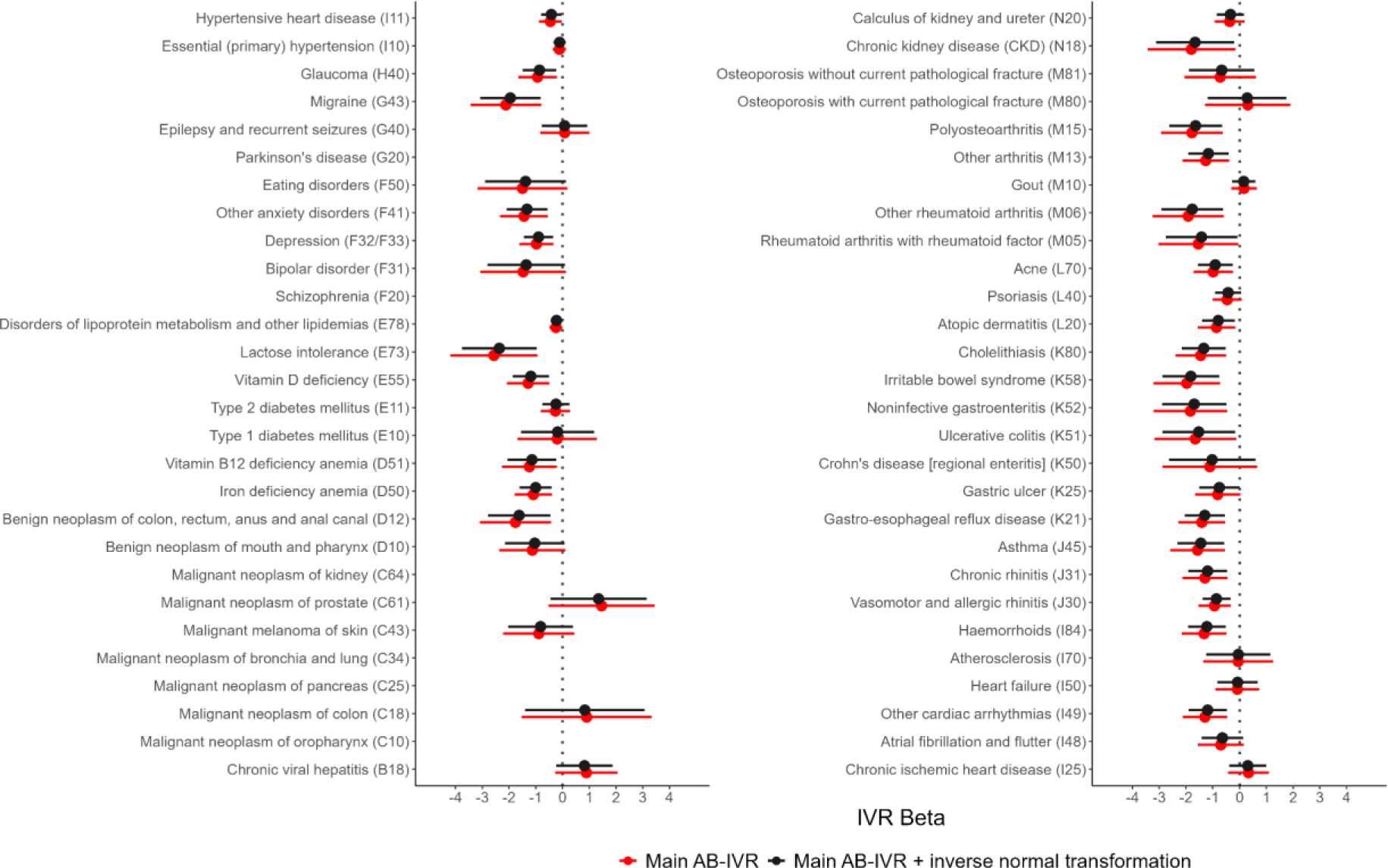
Results of the main analysis for all the 56 studied diseases. Colours represent different data transformations for the *Prevotella/Bacteroides* ratio. Sensitivity analysis results with the inverse normal transformation is shown in black.

**Supplementary figure 2.**
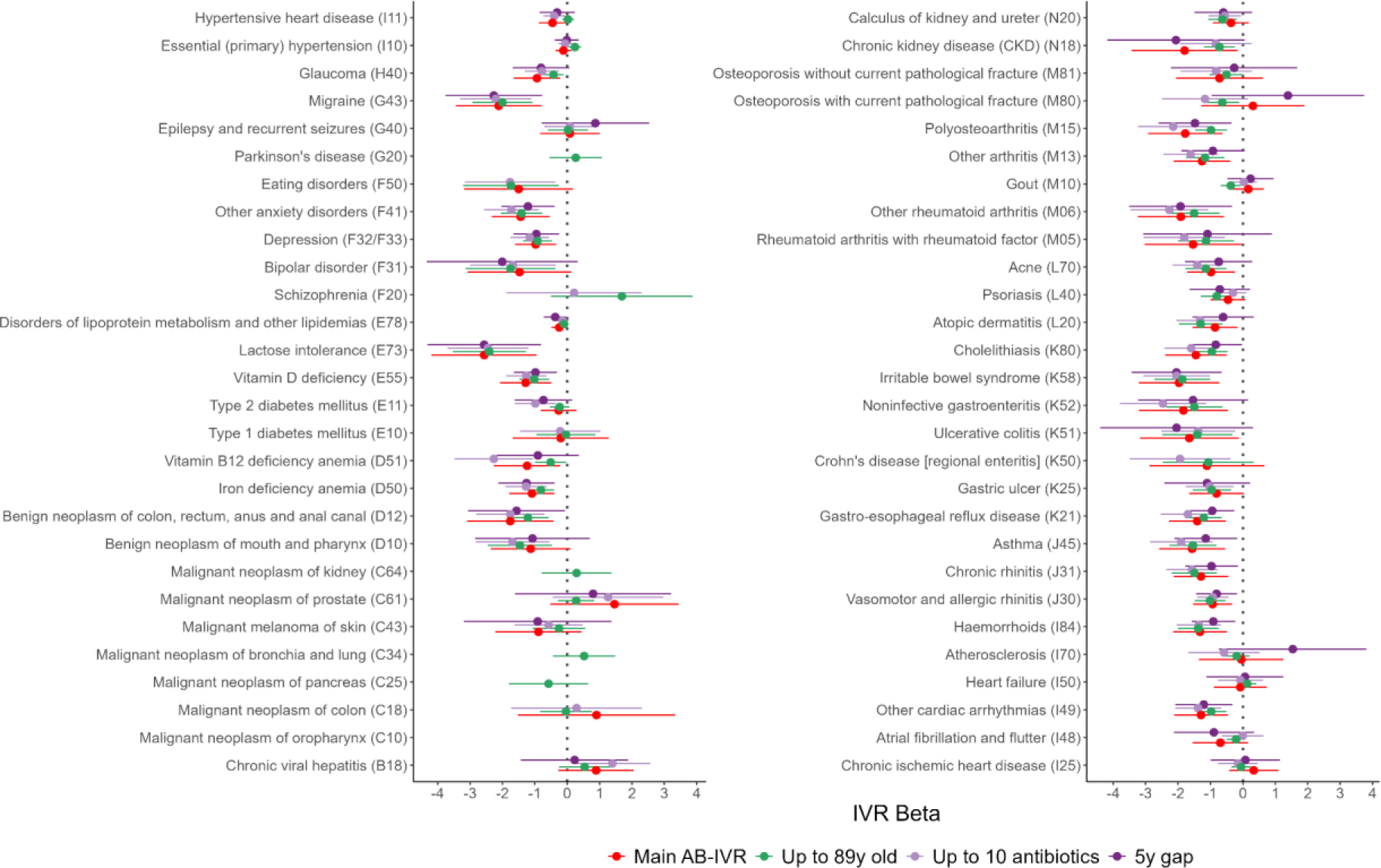
Results of the sensitivity analysis analysis for all the 56 studied diseases. Green shows the effect estimates, when age is filtered as 23-89; light-purple shows the effect estimates, when the maximum number of AB prescribed is 10; dark-purple describes the scenario where the first five years of incidence after the start of follow-up is considered as prevalent disease.

**Supplementary figure 3.**
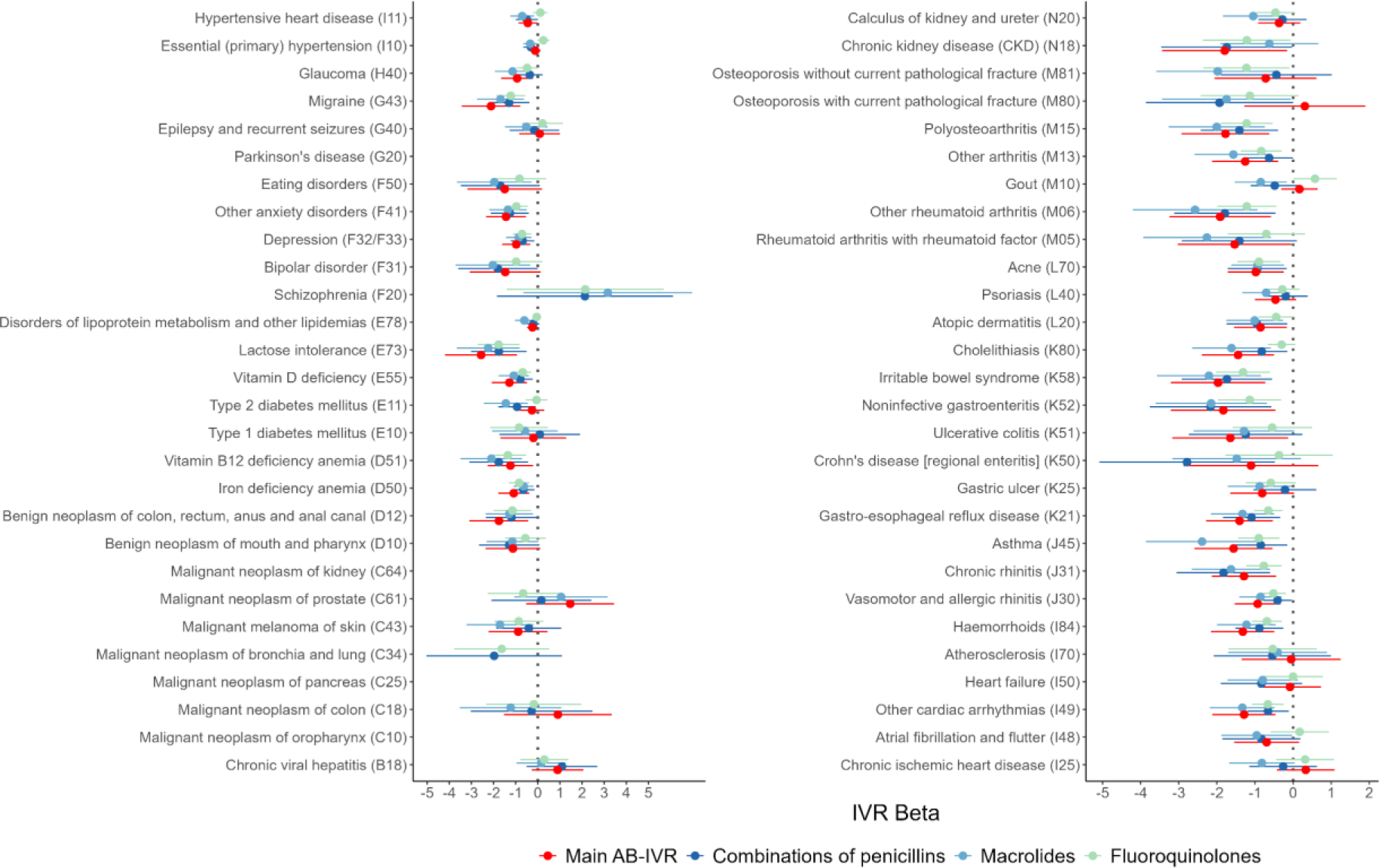
Results of the sensitivity analysis for all the 56 studied diseases. Information regarding subclasses of AB were used instead of the total amount of AB prescribed, whereas other settings were identical to the main analysis. Red corresponds to the main analysis with the antibiotics combined, dark blue corresponds to the class of penicillins (J01CR), light-blue corresponds to the class of macrolides (J01FA) and light-green corresponds to the class of fluoroquinolones (J01MA).

